# Pathological classification of Fuchs endothelial corneal dystrophy in several types and their relationships with CTG18.1 expansion repeats

**DOI:** 10.1101/2025.07.14.25330988

**Authors:** Hanielle Vaitinadapoulé, Daria Onitiu, Corantin Maurin, Gauthier Travers, Emmanuel Crouzet, Oliver Dorado-Cortez, Sylvain Poinard, Zhiguo He, Fabien Forest, Edouard Ollier, Renaud Touraine, Philippe Gain, Jean-Marc Perone, Gilles Thuret, the French Fuchs Study Group (FFSG)

## Abstract

Late-onset Fuchs endothelial corneal dystrophy (FECD) is the most common primary disease of the corneal endothelium and the leading indication for corneal transplantation in Western countries. It is characterized by the abnormal accumulation, over 2 to 3 decades, of extracellular matrix (ECM) components in the form of Descemet’s membrane (DM) excrescents, known as guttae, as well as additional DM layers. Clinical forms and evolutionary profiles vary greatly from patient to patient. It is strongly associated with intronic CTG trinucleotide repeats (TNR) in the transcription factor 4 (*TCF4*) gene. To determine if there are different anatomopathological forms of FECD, we analysed 500 DM removed during keratoplasty for FECD in 25 European centres. After flat mounting and dehydration, the samples were digitized using transmitted light microscopy and observed by 3 independent readers. Ten parameters (6 on guttae and 4 on other forms of ECM) were scored. A principal component analysis and unsupervised clustering method separated 3 clusters from these parameters. In addition, a manual classification, grouping together samples with major common features, isolated 5 types of FECD. The number of TNR in *TCF4* was analysed by ST- and TP-PCR for 109 patients. We found that: 1) 5 phenotypes of FECDs existed, 2) guttae and other ECM structures were radially arranged in 95% of samples, 3) 33% had peripheral radial striae that corresponded to a hypertrophied form of similar structures present in healthy corneas, and 4) the patients with fewer than 50 TNRs had only 2 phenotypes out of 5 and significantly more often peripheral radial striae (94% vs. 49%, P<0.001). Taken together, these new descriptions demonstrate the existence of different FECD phenotypes, reveal that lesions affect both the centre and periphery of the endothelium and suggest that radial deposits may be produced by pathological cells migrating from the periphery to the centre.

## INTRODUCTION

Late onset Fuchs endothelial corneal dystrophy (FECD) is by far the most prevalent primary disease affecting corneal endothelium, representing the first indication for corneal graft in Western countries [1]. It is characterized by quasi-pathognomonic lesions referred to as “guttae”, which are abnormal extracellular matrix (ECM) deposits forming rounded excrescences in the Descemet’s membrane (DM). Guttae gradually accumulate over 2 to 3 decades, in the centre of the DM, forming a growing plaque. In parallel, premature corneal endothelial cell (CEC) death occurs. When the evolution is unfavourable, the endothelial function of regulating stromal hydration becomes insufficient, and permanent corneal edema leads to a significant impairment of visual acuity. Clinical classifications of FECD severity rely on the estimated number of guttae, the diameter of the confluent plaque [2–4] and the presence or not of corneal subedema or edema [5].

The mechanisms involved in premature CECs death and abnormal ECM production are gradually being deciphered and FECD is strongly associated with an intronic CTG trinucleotide repeat (TNR) expansion in the transcription factor 4 (*TCF4*) gene [6], despite significant variations of prevalences (from 76-79% of patients in Caucasians [7], to 26% in Japanese [8]). Several aspects of this mutation remain poorly understood, including understanding if guttae develop differently in patients with and without TNR expansion.

Recently, we described novel structures at the periphery of the DM in FECD, named peripheral striae, appearing as corpuscles with a radial organization resembling pathological long extensions of Hassal-Henle warts [9], and suggested that a dysfunction in the centripetal migration of pathological cells could play a role in FECD pathophysiology [10]. These observations needed to be confirmed in a large cohort.

The heterogeneity of FECD in terms of severity and disease progression is recognized by all clinicians. From a clinical standpoint, this broad spectrum justifies the need to classify each cornea accurately in order to offer personalized care. In the field of basic, translational, and clinical research, this heterogeneity is an obstacle because it limits the development of specific strategies.

To date, the anatomopathological characteristics of FECD have been described and gradually refined over time using mainly corneal cross-sections observed under optical microscopy[11] and transmission electron microscopy [11–13], including serial block face scanning electron microscopy [14]. Flat mounts analysed by scanning electron microscopy have added to our knowledge [15]. To decipher the clinical heterogeneity, we provide here a novel approach of FECD pathology by systematically analysing 500 flat-mounted DMs from FECD patients to identify the types and frequency of ECM lesions and their possible associations. We then propose a new histological classification based on their organizational patterns and compared the genetic characterization of TNR expansion among groups.

## Methods

### Human tissues

The handling of tissues adhered to the tenets of the Declaration of Helsinki of 1975 and its 1983 revision in protecting donor confidentiality and was approved by the ethics committee of the St-Etienne University Hospital (IRB_IORG0007394, Ref_IRBN1142021/CHUSTE). Samples were collected in 25 hospitals (29 surgeons). In this study of surgical specimens, the only clinical data we were authorized to collect were age and sex. Our objective was to analyse DM only, excluding CECs. The only inclusion criterion was a diagnosis of FECD at a stage requiring keratoplasty. We collected the central endothelium on approximately 8 mm in diameter during the first step of the graft, called Descemetorhexis (supplementary material, Figure S1). It was immediately immersed in either balanced salt solution (BSS, Alcon, Rueil-Malmaison, France), or in sterile water to quickly destroy residual CECs, or in 4% paraformaldehyde. Samples were stored at room temperature and sent to the laboratory for analysis.

### Sample preparation and imaging

Each DM was mounted flat on a glass slide (Series 2, Trajan Scientific, Ringwood, Australia) under an operating microscope with the guttae facing upward. They were quickly air-dried and stored in an airtight box at −20°C until observation using transmitted light with bright field illumination (IX81, Olympus, Tokyo, Japan) equipped with a x4 objective and x1.6 zoom and a CMOS camera (ORCA-Flash4.0 LT Digital, Hamamatsu). The mosaic of images (pixel ratio was 0.98461 pixels/µm) was stored in TIFF format. From a collection of approximately 1000 DMs, we selected 545 based exclusively on the quality of the sample (complete disc, if possible, in one piece). We used 500 DMs to establish classifications, and 45 additional DMs to analyse relationships between phenotype and genotype. The collection of 500 images was accessible at the URL: https://gofile.me/5SfuG/6jofA5FOf (password FECD500)

### Classification by systematic analysis of elementary lesions and clustering

We described the characteristic elementary lesions and their distribution over the surface. Based on a training group of 60 DMs, we established 10 simple criteria, achievable without image analysis tools, to describe all possible lesions: 6 for guttae, and 4 for other elements of the ECM (furrows and an embossed appearance of the DM, and structures in the posterior fibrillary layer (PFL)) (**Table 1**). These manual readings were performed by 3 independent readers. Discrepant scores were then reviewed in a concertation meeting to arrive at a consensus score. We verified in terms of agreement between the 3 readers, that we were able to distinguish between small and large diameter guttae or between different areas (for example, more or less than half of the DM area occupied by guttae).

**Table 1.**
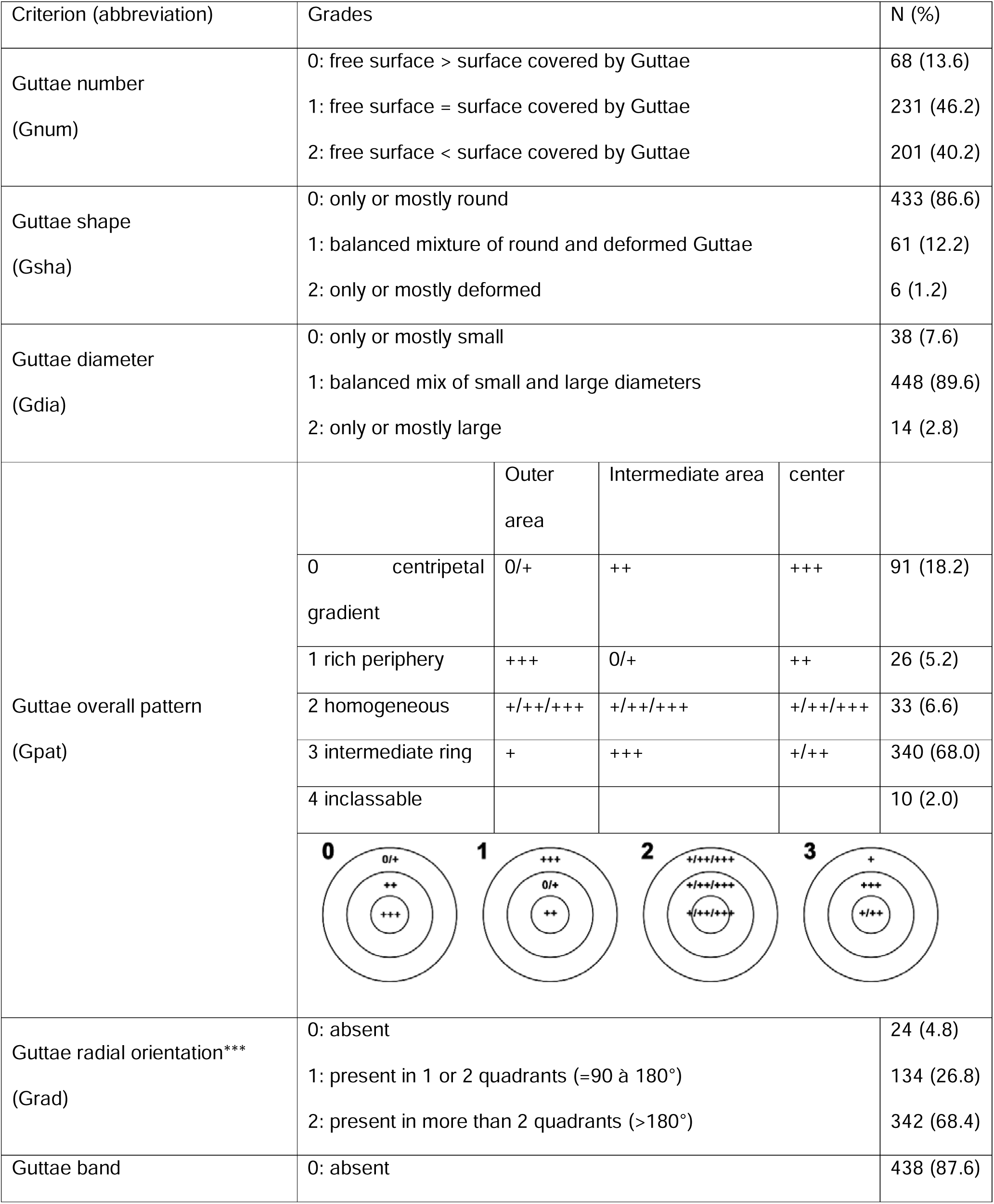
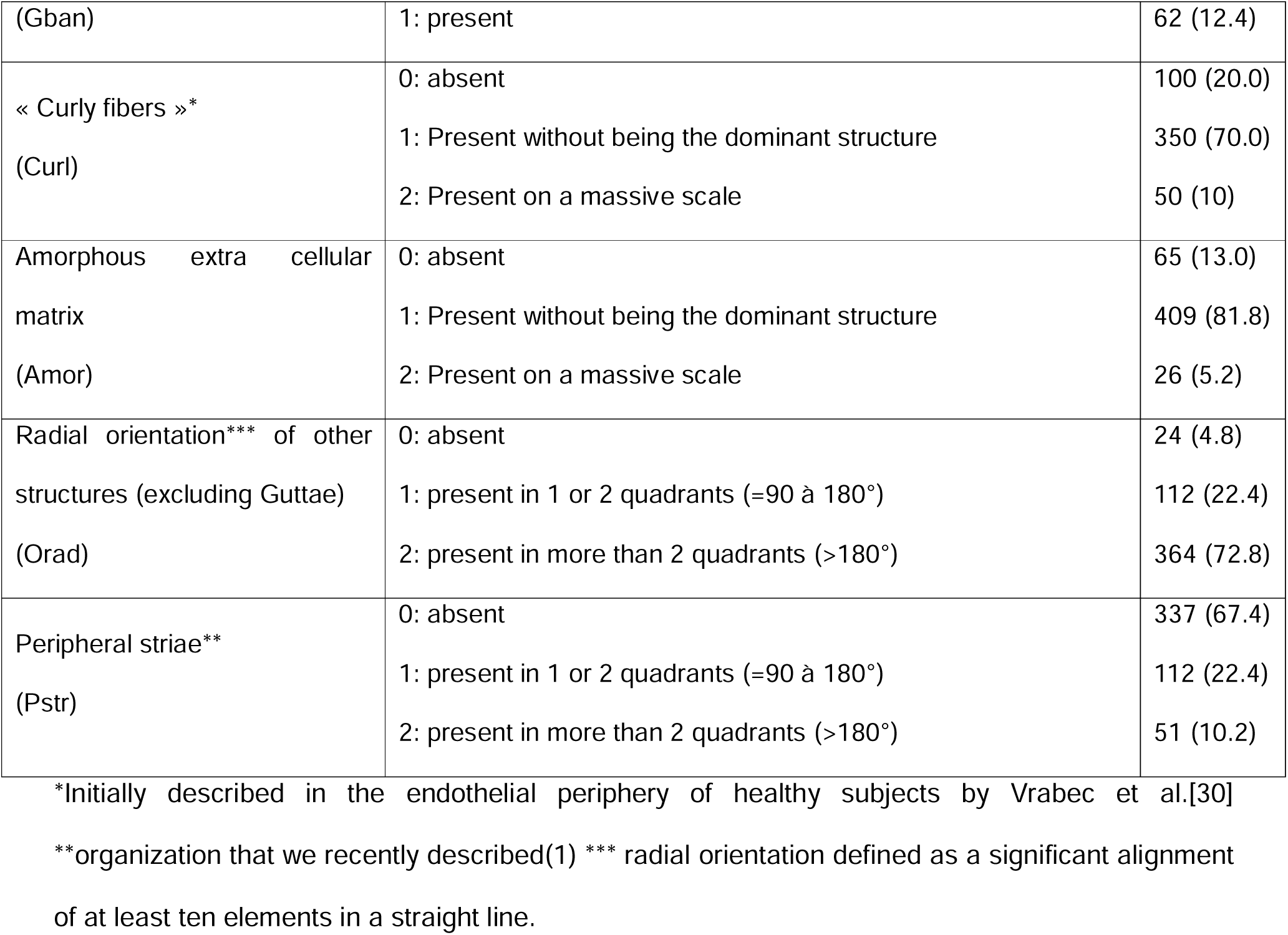
Classification system for each elementary lesion observable on Descemet’s membrane during Fuchs endothelial corneal dystrophy.

We adopted new terminology to describe 3 elements rarely described until now in FECD: 1/ “peripheral striae”, cited above [10], were by definition always at the edge of the sample (interrupted by the surgical tear). 2/ Curly fibres were fibres wrapped around the guttae in varying densities. 3/ “Guttae bands,” defined as very long intertwined bands of guttae with no notable orientation. These 3 structures were illustrated in supplementary material, Figure S2.

Three parameters were quantified by image analysis (FIJI, https://imagej.net/software/fiji/downloads). We measured the minimum and maximum Feret diameters of each DM in order to analyse whether the frequency of peripheral striae located at the edge of the sample was underestimated in the smallest ones. We also measured the diameter of the guttae on a random sample of DMs (7 in each of the 3 categories of guttae diameter). We manually measured the maximum possible number of guttae within a 200 µm wide band, running from one edge to the other, passing through the centre, using the straight-line selection in Fiji software.

### Manual classifications

After noting significant similarities and differences the DMs, we also carried out a manual classification, establishing two rules: identify a single dominant characteristic as often as possible; if this is not possible, choose a maximum of two or three characteristics. This classification was carried out by two observers (HV and GT). The groups and their intersections were represented using a proportional Venn diagram [16], programmed in R (https://www.r-project.org).

### Genotyping

To investigate histological differences between TNR expansion-positive and TNR expansion-negative samples, we simultaneously characterized the genetic profiles and the histological phenotypes for 109 patients selected at random, including 64 patients from the series of 500 classified DMs. Details about the two PCR used were exposed in the **Supporting Information.**

### Statistical analysis

Data were described by their mean +/- standard deviation (SD), median (min-max). Data with a normal distribution were compared using parametric tests. Otherwise, a non-parametric tests were used. P<0.05 was considered the significance threshold unless otherwise stated. We performed principal component analysis (PCA) on the 10 manual classification variables, using the oblimin rotation method, as we assumed that residual correlations might exist between the principal components (PCs).

Two-step clustering analysis, i.e., pre-clustering followed by hierarchical clustering, was applied on the 10 variables of the manual classification to automatically identify the optimal clustering number based on the silhouette width. The distance was calculated using the log-likelihood and clustering by Schwarz’s Bayesian criterion.

Statistics and graphs were performed using IBM SPSS Statistics software. 30.0.0.0. except for the violin plots (generated on the website https://www.statskingdom.com).

## RESULTS

### General characteristics and influence of preanalytical conditions

Each centre collected 21+/-24 DMs (median 14, from 2 to 99). Patients age was 70±9 years (median 71, range 41 to 95 years) and did not differ significantly between centres (P=0.331). They were 66.4% women and 31.4% men (2.2% missing data). Ages of men and women did not differ (p=0.741). The minimum and maximum ferret diameters were 8.3+/-1.1 mm and 9.5+/-1.1 mm, respectively. The influence of the 3 transport liquids on the grading of elementary lesions was detailed in the supplementary material, Figure S3, and can be summarised as follows: we did not note any morphological changes in the various MEC abnormalities at the level of our observations, but for three of the ten items (Amor, Grad, Orad), the MDs in PFA had lower scores, suggesting that the fixed residual cells interfered with the observation of the finest structures (masking effect).

### Elementary lesions: descriptive data

Frequencies distribution of individual lesion was detailed **Table 1** and could be summarized as follows: the number of guttae was significant or very significant in 86.4% of DMs (grades 1 and 2 of item Gnum). They were predominantly round (86.6%) and in 89.6% of cases presented a balanced combination of small and large guttae (grade 1 of item Gdia). The majority (68.0%) of DMs showed a pattern with a central area of guttae surrounded by an even denser ring of guttae and then a much less dense area of guttae (grade 3 of the Gpat item).

We identified 3 structures which had a radial organization (**Figure 1A**): i/ In 95.2% of DMs, guttae (regardless of their diameter) were perfectly aligned, forming radially oriented lines that could be present over a variable surface area, limited to 1 or 2 quadrants in 26.8% of cases and extending to 3 or 4 quadrants in 68.4% of cases. These aligned guttae could coexist with randomly distributed ones. ii/ two structures (furrows or “embossment” in the DM) also adopted a radial alignment in 95.2% of DMs with a variable extent over 360°. They were significantly associated with radial guttae (Spearman’s coefficient = 0.255, P<0.001). iii/ Peripheral striae were present in 32.6% of cases. They were found around 360°, though they were sometimes visible in only one, two, three, or all four quadrants (possibly depending on the centration of the surgical dissection). Samples without peripheral striae were significantly smaller (min and max Feret diameters, 8.04+/- 1.01 and 9.24+/-1.00 versus 8.86+/-1.06 and 10.19+/-0.97, P<0.001 for both diameters) than those with striae, suggesting that we may have underestimated the percentage of striae on DMs that were small in diameter. There was no significant association between the radial organization of the guttae and the presence of peripheral striae (Khi^2^, P=0.973).

**Figure 1.**
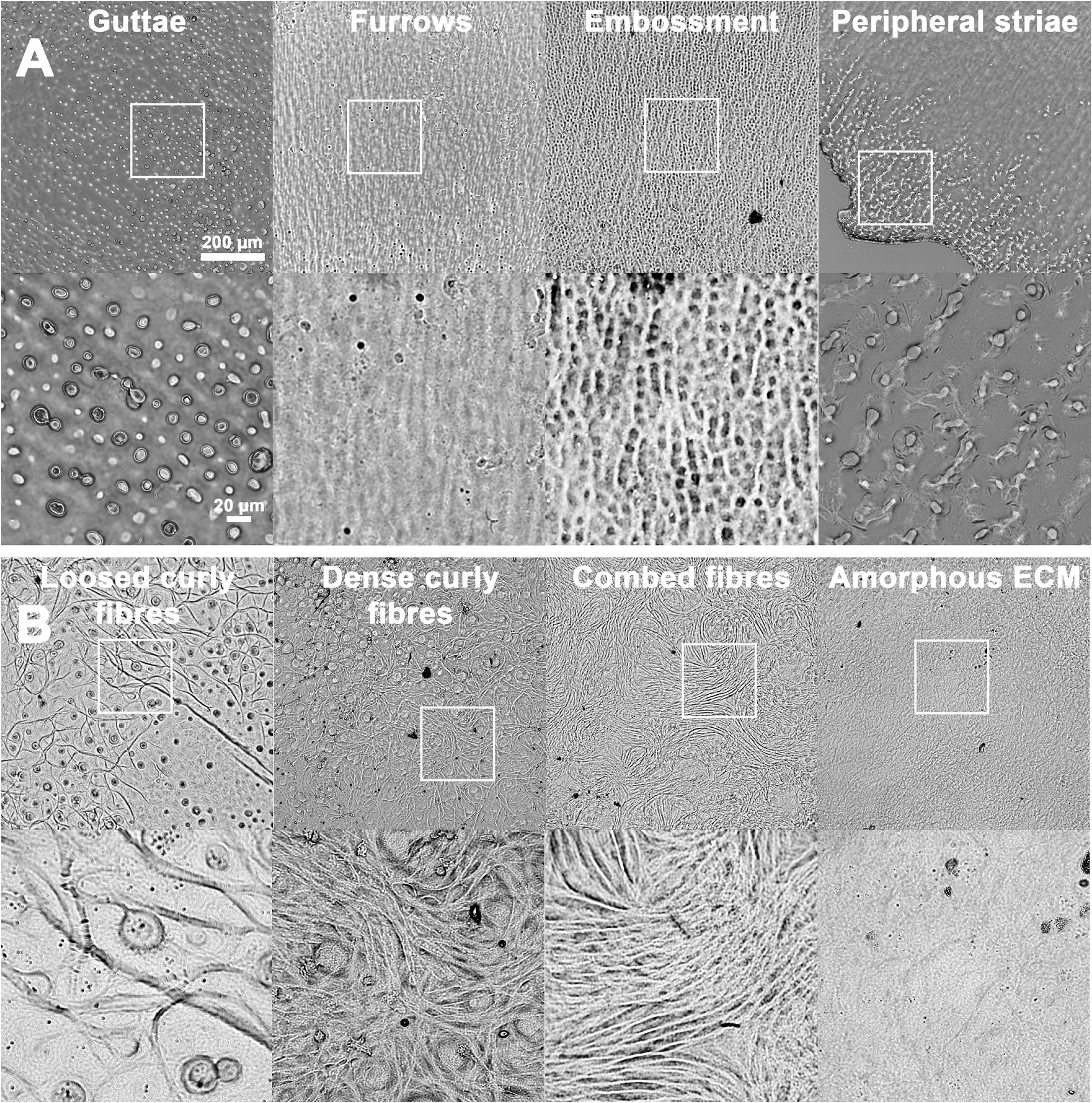
Structure of the guttae, Descemet’s membrane furrows and embossing, peripheral striae and posterior fibrillar layer (PFL) at low and high magnifications (x10 and x40 objectives). (A) The four radially organized structures. (B) The different structures composing the PFL.

We also identified two other ECM abnormalities (**Figure 1B**) that were located mainly (but not exclusively, as shown by the existence of a group of outliers) in the centre of the DM, above the guttae that often remained visible through transparency: 1/ curly fibres, with more or less tight loops, were present in 80.0% of cases and were the dominant structure (in terms of surface area) in 10.0%. More rarely they appeared as combed fibres. 2/ A layer of amorphous ECM covering the guttae was observed in 87.0% of cases and was the dominant structure in only 5.2% of cases. Only 5.8% (29/500) of DMs had neither of these two forms of abnormal ECM.

### Elementary lesions: distribution of the diameter of the Guttae

The diameters, measured on 15841 guttae, were 10.5+/-4.12, median 9.93 (3.0-37.2), 16.6+/-5.9, median 15.4 (6.4-67.6) and 21.9+/-6.6 median 21.3 (7.5-63.5) µm for the small, mixed and large guttae groups respectively (p<0.001). Two-by-two tests showed significant differences (Bonferroni correction) (**Figure 2)**. This quantitative result indicated that our subjective classification of diameters effectively separated the 3 groups, and that the mixed group seemed to correspond to guttae of intermediate diameter rather than to 2 distinct populations. In all groups, the diameter range was very wide, confirming the diversity of intra- and inter-patient diameters.

**Figure 2.**
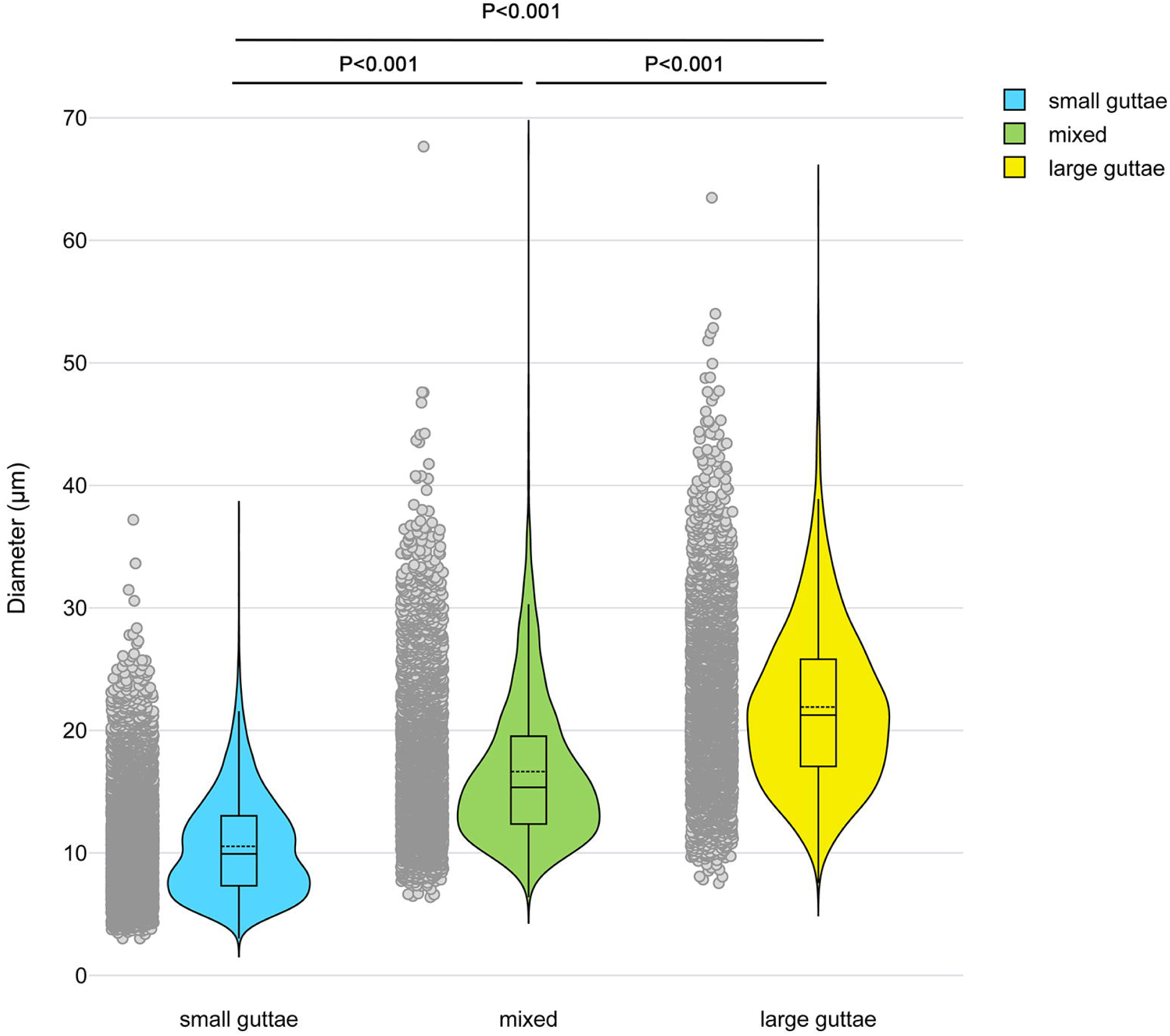
Guttae diameter. Scatter, violin and box plots of the 3 categories of Guttae diameter separated by our *manual* classification.

### Histological phenotypes by principal component analysis and clustering

Four main components (PCs) were obtained, explaining 59% of the total variance. PC1 corresponded broadly to the characteristics of the ECM, PC2 to the radial nature of the different structures, and PCs 3 and 4 to the characteristics of the Guttae. PCs 1 and 2 were shown in **Figure 3**.

**Figure 3.**
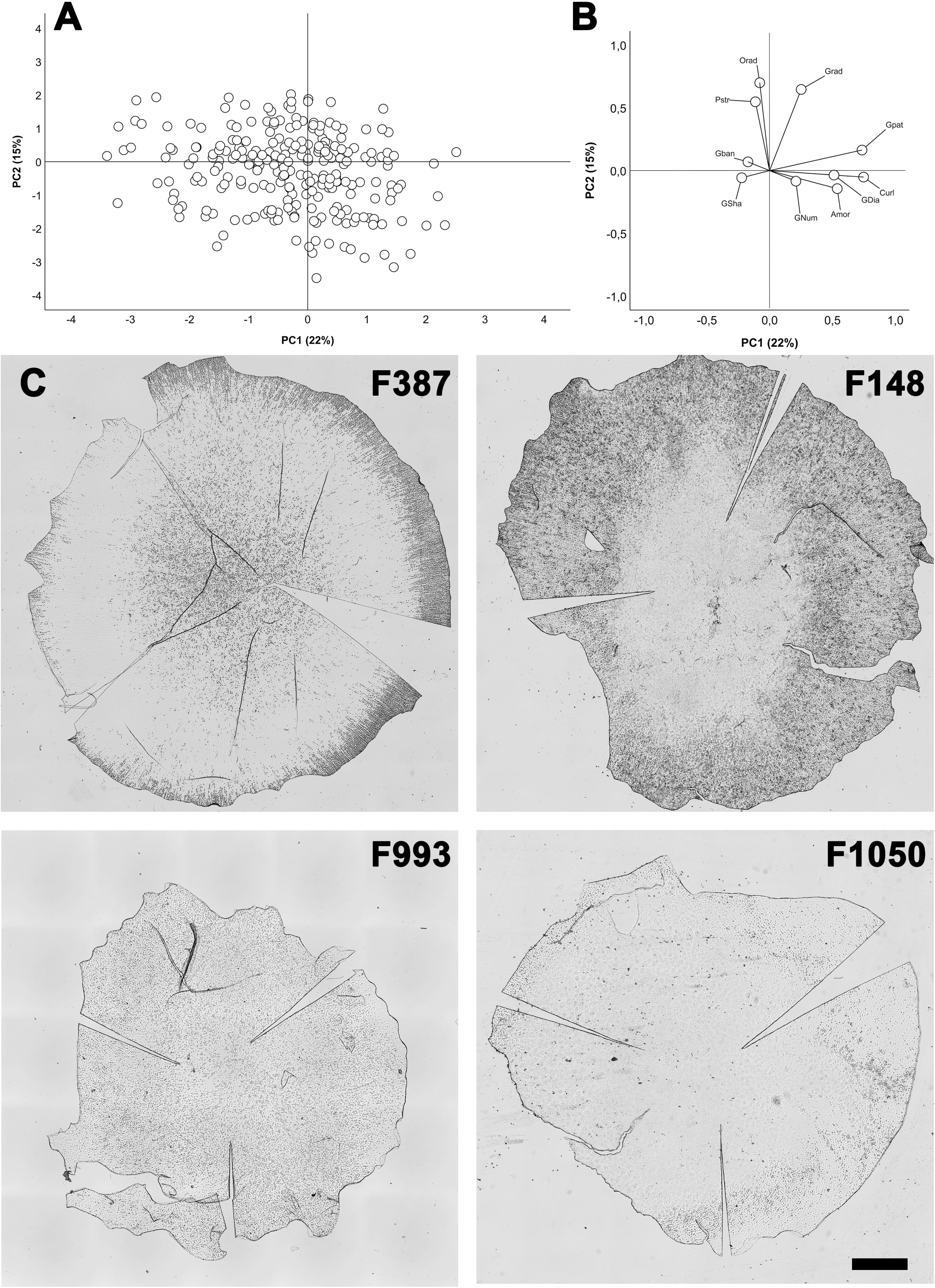
Graphical representation of the results of principal component analysis. (A) Projection of samples in the space constructed from the first two principal components (PC). (B) Contribution of the 10 variables to each of the first two PCs. Gpat: Global guttae pattern; Gnum: Guttae number; Gsha: Guttae shape; Gdia: Guttae diameter; Curl: Curly ECM; Amor: Amorphous ECM; Grad: Radially aligned guttae; Orad: Other radially aligned structures; Pstr: Peripheral radial striae; Gban: Bands of guttae. (C) Example of four Descemet membranes located at the extremes of the factorial planes of the first two PC. The references allowed them to be found in the online image collection to view them in full resolution. F387 is notable for the presence of peripheral radial striae, F148 is notable for the large elliptical center of confluent guttae surrounded by aligned and radial guttae, F993 is notable for the presence of only small, round guttae, and F1015 is notable for the almost complete coverage of curly structures that mask the guttae. Scale bar 1 mm.

Employing two-step clustering revealed 3 distinct clusters, as depicted in **Figure 4** and **Table 2**. The clustering effect was nevertheless weak (silhouette[=[0.2), and the importance of predictive factors was variable. Cluster 1, representing radial organization (211 cases, 42%), encompassed DM with high scores in radial guttae and other structures and a moderate score in peripheral striae which are also by definition radially organized. It was dominated by an overall organization with a confluent centre surrounded by a ring of dense guttae then by a periphery with few Guttae (sub-type 3 in **Table 1**). Cluster 2, (116 cases, 23%) resembled cluster 1, but had less numerous guttae, smaller ones and less often central ECM. Cluster 3 (173 cases, 35%) had the larger amount of guttae of all sizes and was dominated by large plaques of curly and amorphous fibres covering central confluent guttae. We found no difference between clusters 1, 2, and 3 in terms of sex ratio, with 69%, 65%, and 69% of women, respectively (Khi^2^, P=0.780), but there was a significant difference in age, with 70+/-8 (46-91), 73+/-10 (42-93), and 70+/-10 (41-95) years old (P=0.002, Kruskal-Wallis + Dunn’s post hoc), and cluster 2 being significantly older than the other two.

**Figure 4.**
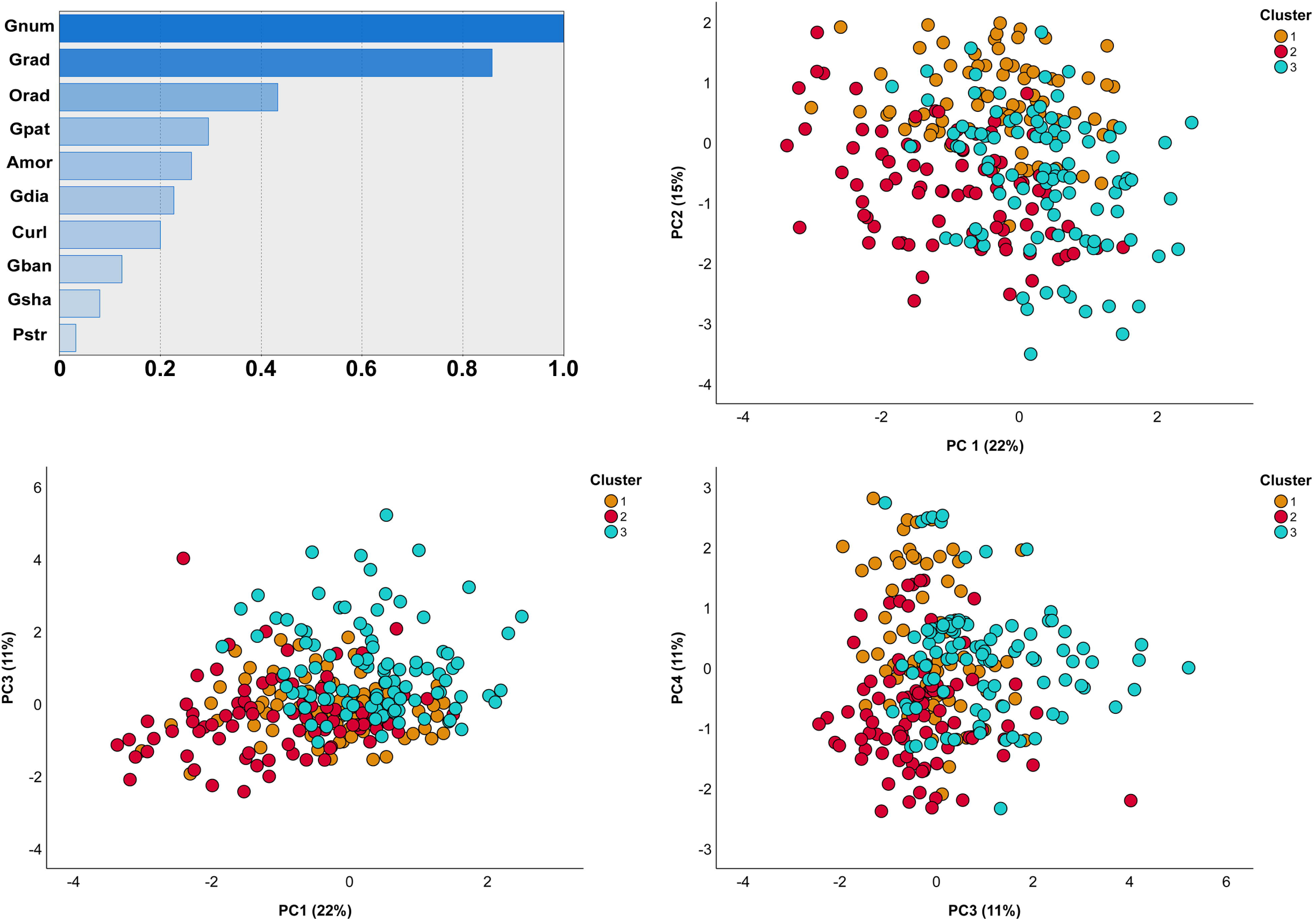
Contribution of the histological components to the clustering solution as reported from the Two-Step procedure. The top left panel showed the index of relative importance of each histological component as identified by the Two-Step cluster analysis. Representations of the 3 clusters in two dimensions using the principal component analysis. We chose to represent 3 graphs showing the partial separation of the clusters whatever the dimensions chosen.

**Table 2.**
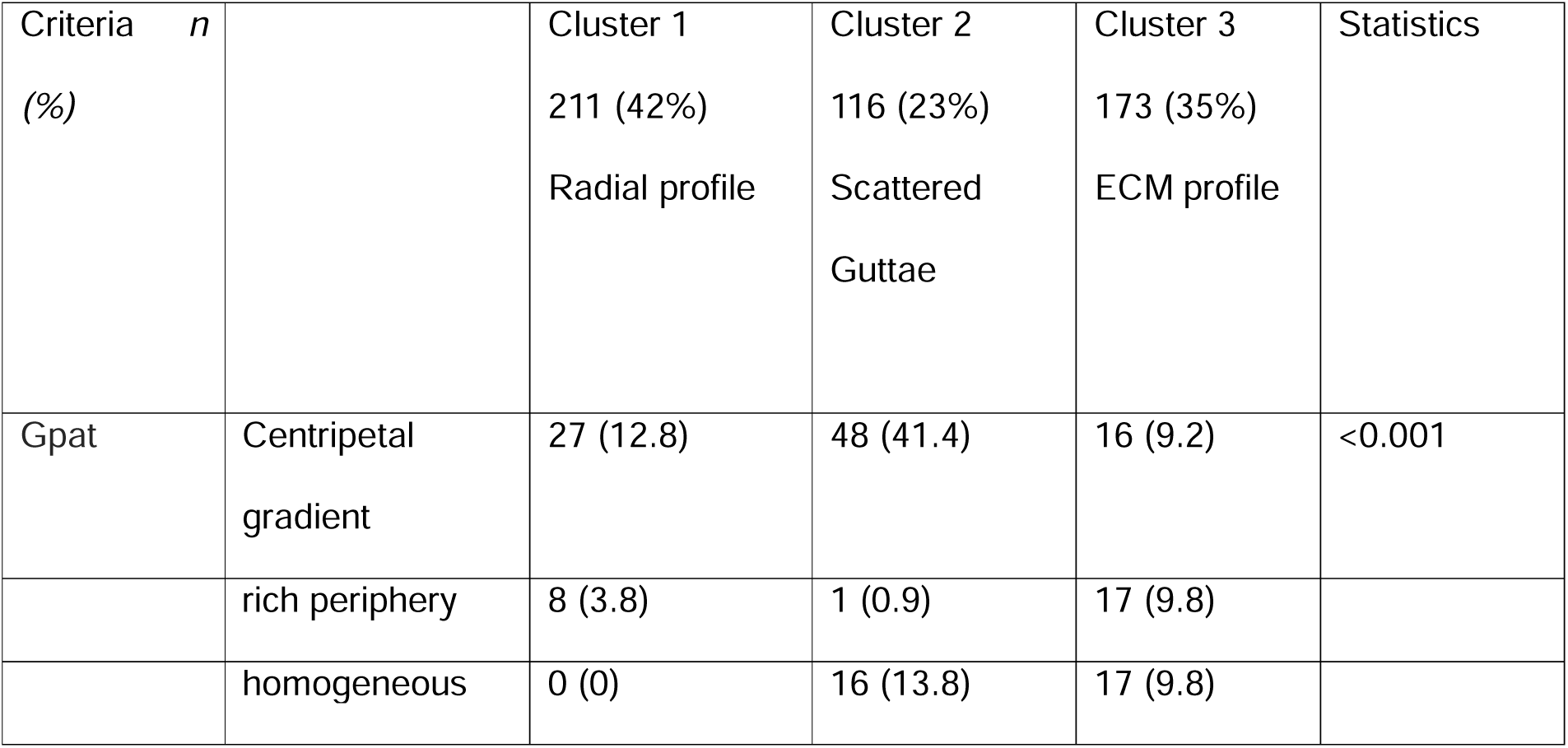
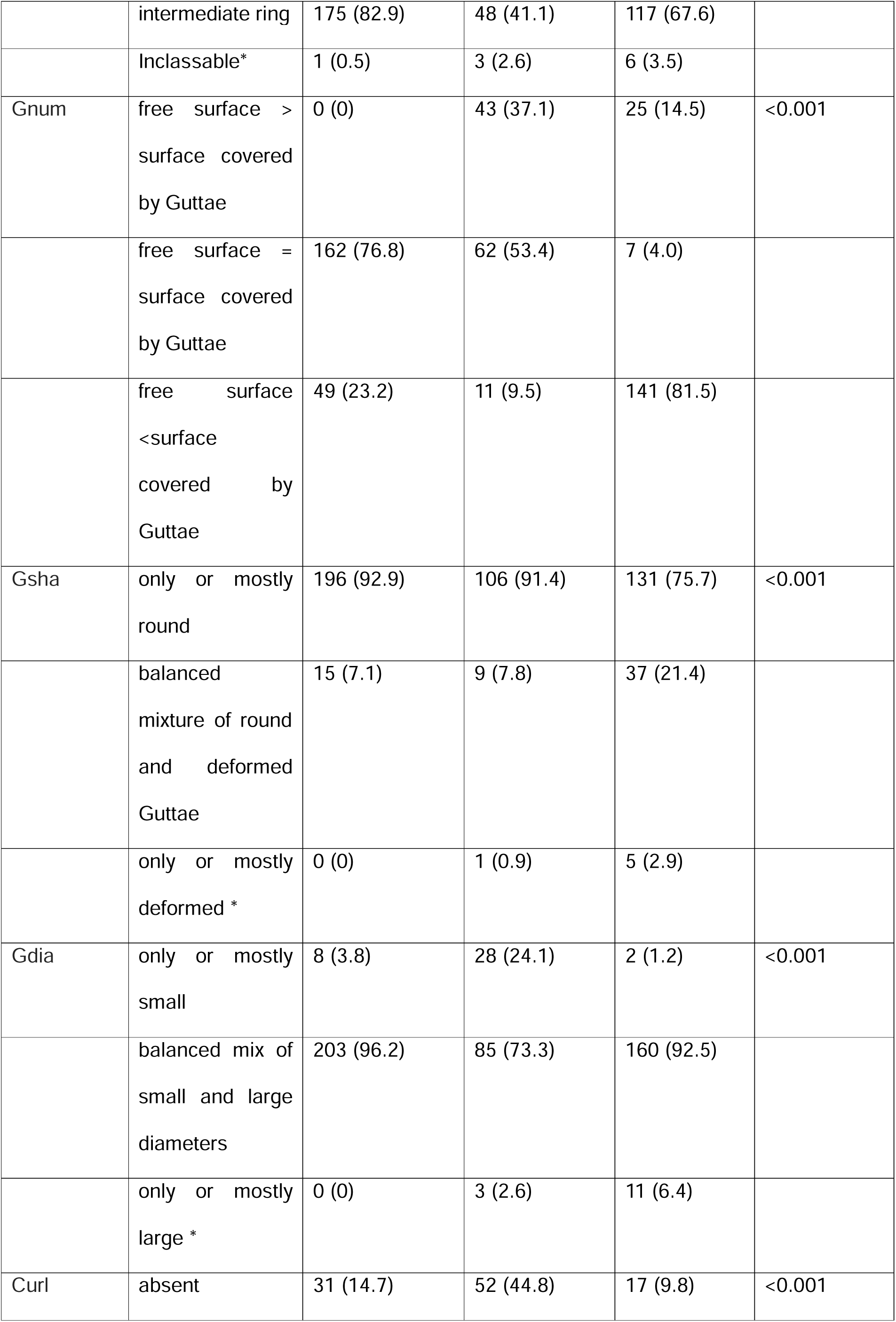
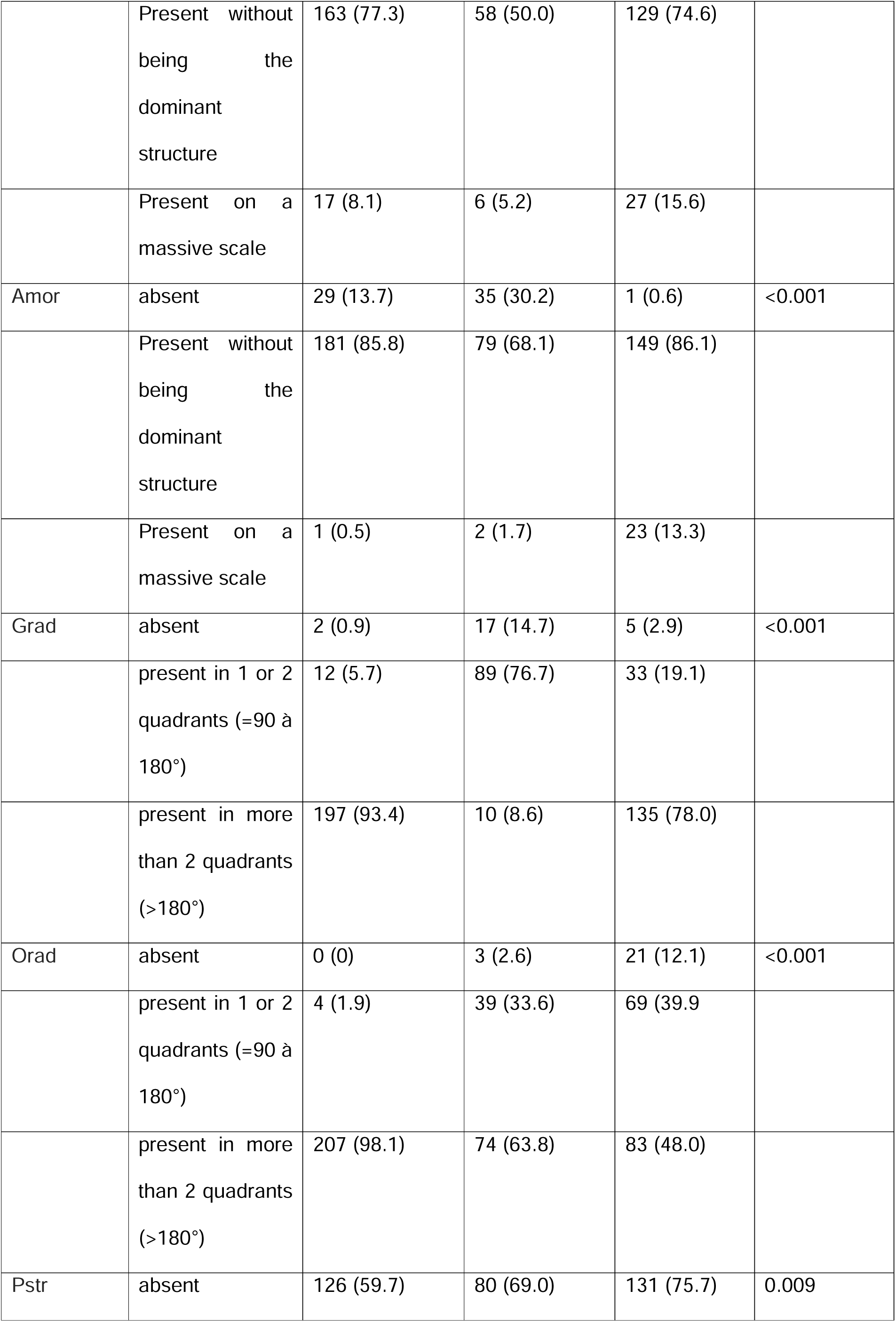
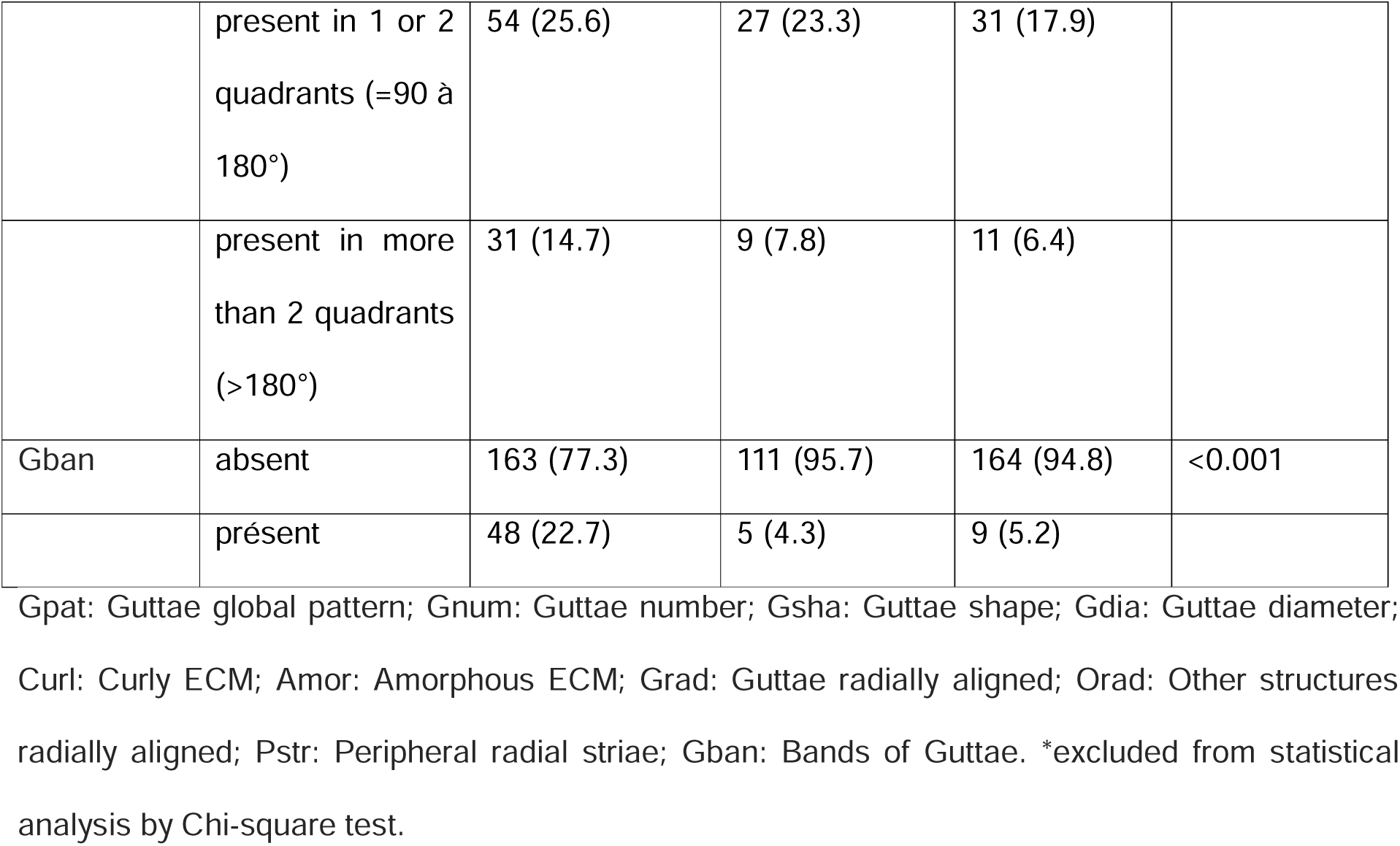
Histological characteristics in the 3 clusters extracted by the two-step clustering method.

### Histological phenotypes by manual classification

We isolated five main groups or dominant phenotypes: 1/ round or elliptical central structure where the guttae were covered with ECM (corresponding to the conventional conception of FECD): single Centre group for 54.0% of cases; 2/ organization in radial lines (an organization that we recently described[10]) : Radial group for 27.6% of cases; 3/ organization in several epicentres: epicentre group; 4/ presence of guttae only without added ECM, without a clearly organized central zone: Guttae-only group for 9.4% of cases. It was the most isolated group, with only 7 cases in common with the Radial group and 1 case with the Fused group.; 5/ fusion of guttae: Fused group. It was the least independent group, with only 2 “pure” cases and 26 cases sharing other morphological characteristics. Only 34 DMs (6.8%) could not be classified. For the remaining 466, the groups were presented in **Figure 5**. Eighty percent were considered to have a single dominant feature, and 20% showed significant overlap between groups. Given this overlap, the age and sex distribution were expressed in **Table 3** in two ways: for all patients in the 5 groups (with some patients counted 2 or 3 times) and for the 5 subgroups of patients with a unique characteristic. Regardless of the grouping method, we did not find any significant differences in mean age or sex ratio between the five phenotypes.

**Figure 5.**
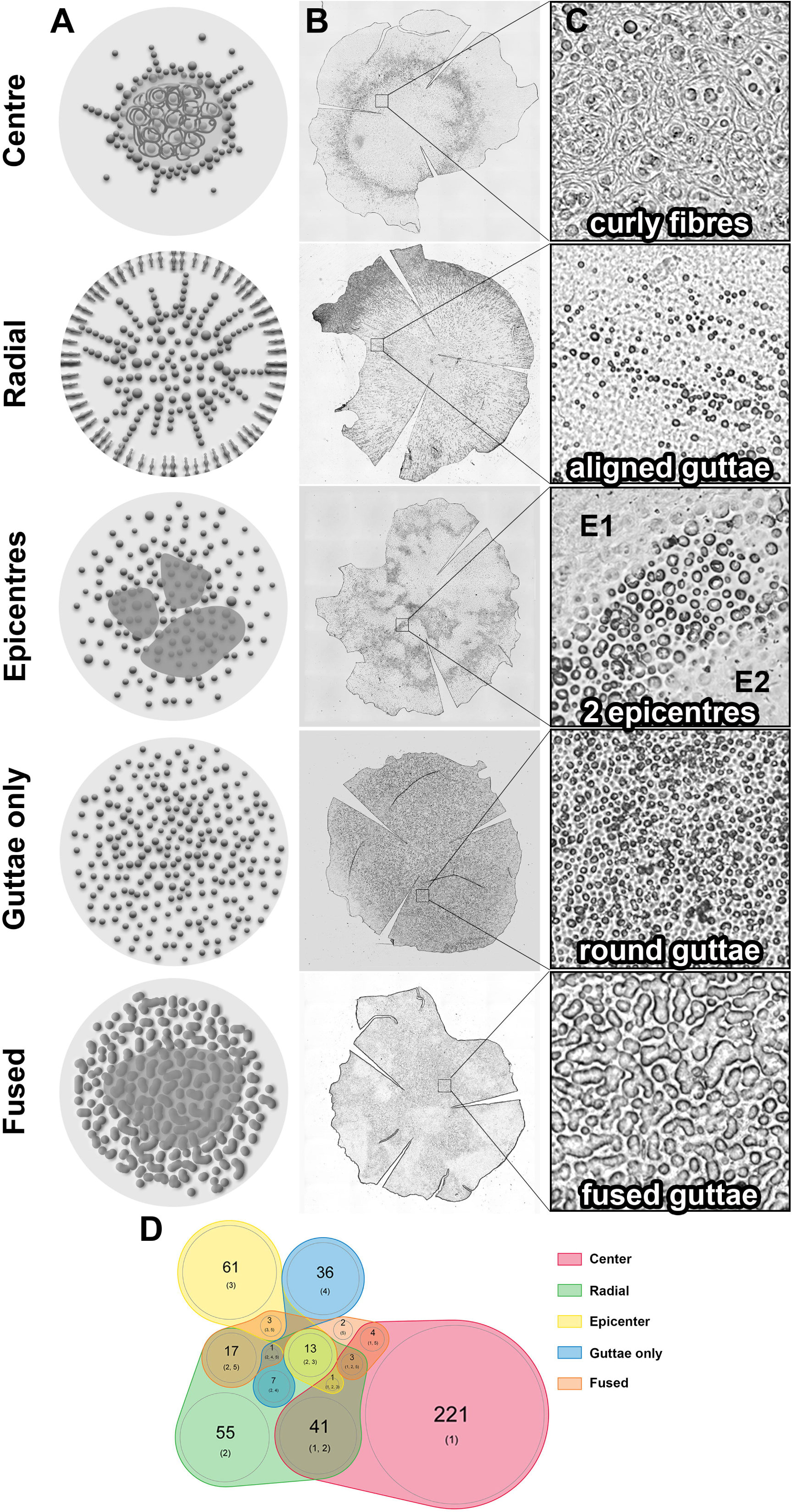
Five histological phenotypes of Fuchs endothelial corneal dystrophy identified by manual classification. (A) Schematic representation of the distribution of histological lesions in the central 8 mm of the endothelium (red circle). (B) Descemet’s membrane of FECD patients for each of the five phenotypes (low magnification X6.4): phenotype 1 (single center with curly fibers and guttae ring), phenotype 2 (radial distribution with striae and bands of guttae), phenotype 3 (epicenters with curly fibers spots), phenotype 4 (round guttae only) and phenotype 5 (central fused guttae and peripheral round guttae). (C) High magnification (X40) of lesions characteristic of each histological phenotype. The images were obtained by transmitted light microscopy. (D) Proportional Venn diagram of our proposed histological classification.

**Table 3.** Age and gender distribution among the 5 phenotype groups.

| Type of analysis | Phenotype | N (%) | Age (year) mean (SD) | Sex (% female) |
| --- | --- | --- | --- | --- |
| All cases (n=581) | Center | 263 (45.3) | 70.7 (8.6) | 68.8 |
|  | Radial | 134 (23.0) | 69.8 (9.3) | 77.6 |
|  | Epicenter | 77 (13.3) | 68.2 (7.9) | 61.0 |
|  | Guttæ only | 43 (7.4) | 71.9 (10.1) | 65.1 |
|  | Fused | 30 (5.1) | 66.8 (10.7) | 83.3 |
|  | Outlier | 34 (5.8) | 72.0 (11.0) | 61.8 |
|  | P |  | 0.010 (Kruskal-Wallis) | 0.053 (Khi<sup>2</sup>) |
| Only cases with unique characteristics (n=403) | Center | 218 (54.1) | 71.9 (8.6) | 65.0 |
|  | Radial | 53 (13.2) | 72.4 (9.0) | 73.6 |
|  | Epicenter | 61 (15.1) | 68.6 (7.7) | 58.3 |
|  | Guttæ only | 35 (8.7) | 71.9 (10.7) | 68.6 |
|  | Fused | 2 (0.5) | 66.0 (8.5) | 50.0* |
|  | Outlier | 34 (8.4) | 72.0 (11.0) | 61.8 |
|  | P |  | 0.066 (Kruskal-Wallis) | 0.518 (Khi<sup>2</sup>) |

\* excluded from statistical analysis due to insufficient sample size.

### Relationship between phenotype and triplet repeats in the TCF4 gene

Of the 109 FECD patients genotyped, 84.4% had TNR expansion. The distribution of homozygotes and heterozygotes was detailed in **Table 4**. The distribution of histological phenotypes according to the TNR expansion was detailed in **Figure 6**. The phenotypes of the two groups were significantly different (Chi-square test with Yates correction, P<0.001). Notably, for the 17 DMs from patients without TNR expansion, the phenotypes included only radial and/or fused components. In addition, they significantly more often presented peripheral striae than those with TNR expansion: 16/17 (94%) versus 45/92 (49%) (Chi-square test, p<0.001). The striae were also more extensive over 360°: 10/16 (63%) versus 10/45 (22%) (Chi-square test, P<0.001). Conversely, no patients with TNR expansion had fused guttae.

**Figure 6.**
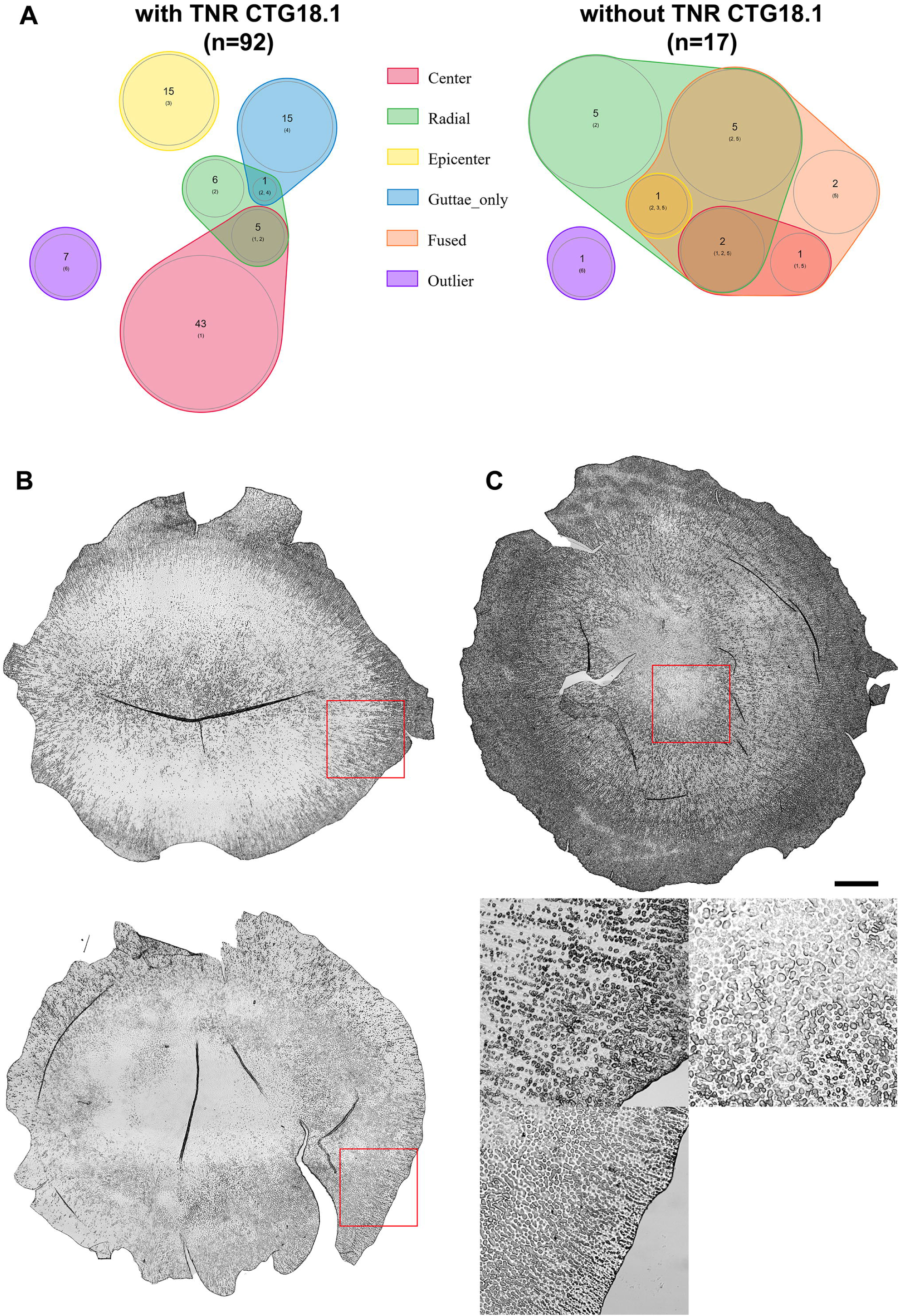
**Distribution of phenotypes according to the presence or absence of CTG 18.1 triplet repeat (TNR) in the *TCF4* gene**. (A) The distribution was different between the two groups as shown by the Venn diagrams (P<0.001). Three prototypical examples of pathological phenotypes predominant in patients without CTG18.1 TNR. (B) 84-year-old woman presenting with radial organization and peripheral striae over 360°. (C) 76-year-old woman presenting with a combination of radial + fused guttae and extremely dense peripheral striae over 360°. (D) 59-year-old woman presenting fused guttae and peripheral striae. Scale bar 1 mm. Bottom right, 1 mm^2^ zoom on characteristic lesions: fused guttae and peripheral striae.

**Table 4.** CTG18.1 Trinucleotide repeats length in *TCF4* gene among FECD patients.

| Number of repeats |  |  | Homozygote | Heterozygote | Total | % |
| --- | --- | --- | --- | --- | --- | --- |
| Expansion-positive | >50 | <100 | 6 | 27 | 33 | 30,3 |
|  |  | >100 | 3 | 56 | 59 | 54,1 |
| Intermediate | 40-50 |  | 0 | 2 | 2 | 1.8 |
| Expansion-negative | <40* |  | 2 | 13 | 15 | 13.7 |

\* of which 11 with less than 32 TNR to comply with the RNA focus formation threshold according to
Zarouchlioti et al.[39]

## Discussion

The high prevalence of late-onset FECD in people over 40, its increase linked to aging [17], and new short- and medium-term therapeutic prospects will require new diagnostic criteria to personalize care. The existence of a diversity of clinical forms and evolutionary profiles, observed by all clinicians and debated [18], has not, to our knowledge, yet been linked to different histological forms. Based on a large series of cases we analyse the elementary lesions characteristic of FECD and propose a new classification.

FECD is considered a disease of the central endothelium that gradually spreads toward the periphery through the accumulation of guttae and premature cell death. The historical classification by Krachmer, which is still the most widely cited [19], defines six stages of increasing severity but do not separate different clinical and/or histological forms. It is based on the number of guttae and the area of confluent guttae, and places the onset of edema in the last stage when the confluent plaque exceeds 5 mm in diameter [2]. However, some corneas present edema without having such a large confluent area [20]. We show figure 5 that the radial phenotype rarely presents confluent guttae and that the guttae-only phenotype never does.

Our series is representative of the conventional understanding of FECD at the surgical stage: it includes mostly women, patients were most often operated on between the 6th and 7th decades, and the majority presented with very numerous guttae, confluent in the centre. However, our systematic analysis reveals, for the first time to our knowledge, other histological forms that differ from this usual prototype.

The diameter of guttae is the first element that allows different groups to be defined, as we had already assumed by measuring the diameter and height of 3 different guttae types using chromatic confocal microscopy [21]. It is generally accepted that guttae diameter increases over time as severity increases [22]. The different types of guttae we have identified suggest significant variations in their formation and growth mechanisms: while the majority (90%) of DMs contain guttae of different diameters, suggesting different ages, DMs from the guttae only group contain tens of thousands of guttae of virtually identical small diameters, consistent with the hypothesis of synchronized development and showing that severe forms of FECD exist without large guttae.

We highlight a very high frequency of ECM elements with a radial arrangement which we already described clinically [10, 23]. Peripheral striae constitute a new structure in FECD. They have been overlooked until now probably for 2 reasons: 1/ the endothelial periphery is very difficult to visualize with a slit lamp, often obscured by the arcus senilis, and non-contact specular microscopy is limited to the central 6 mm; 2/ they are invisible on conventional cross section histology. The peripheral striae appeared to be pathological extensions of the shorter striae we described at the extreme periphery of healthy corneas from elderly donors [9]. Our work clearly demonstrates that FECD is not limited to the centre of the endothelium.

We assume that all these linear and radial structures could be traces left in the DM during cell migration. Normal and diseased CECs secrete ECM proteins, metalloproteases and their endogenous tissue inhibitors, which modulate the tissue [24]. Different cells could produce the different structures: normal CECs for DM structures that are not very prominent (embossments that may correspond to the location of the cell nucleus) and furrows that also exist in healthy cornea donors [9] and abnormal CECs responsible for the formation of guttae and peripheral striae along their path. In this context, two recent concepts should be considered: the existence of several subpopulations of CECs [25, 26] and a possible increase in the in vitro migration speed of FECD CECs, particularly in cases with TNR expansion [27, 28]. We assume that this migration is centripetal since these alignments are found both in the centre and at the periphery and since it is also consistent with embryology (cells migrate from the periphery of the presumptive cornea to the centre) [29]. In addition, the frequent “epicentre” phenotype could be explained by an heterogeneous distribution of abnormal peripheral CECs and/or cells which do not migrate exactly toward the centre of the cornea or not at the same speed.

Our work provides new insights into the structure of PFL, which can comprise two types of ECM. It is either highly organized in the form of curly or combed fibres forming a network around and above the guttae, or amorphous, masking the buried guttae. The two forms of ECM can coexist. Note that the term “curly structure” was first used in 1974 to describe a similar organization located between Hassal-Henle bodies at the extreme periphery of healthy endothelium [30]. The existence of these two forms of ECM proteins could correspond to different isoforms, by homology with the different deposits of transforming growth factor-β-induced protein mutated in epithelial-stromal dystrophies [31]. In addition, we have just described that curly fibres contain specific proteins (Tenascin-C and Byglican) [32, 33]. Further analysis will be needed to compare proteins in the two types of ECM.

We were struck by the similarities between the periphery of healthy endothelium and the centre of certain FECD groups (centre, radial, epicentre): both have excrescences in the DM (Hassal Henle warts and guttae), striae [9] and curly fibres or structures[30, 34]. Their common feature is the presence of poorly differentiated CECs in the healthy periphery (progenitor) [9, 35] and dedifferentiated cells in the centre through endothelium-mesenchymal transition mechanisms in FECD [36].

Our two classifications do not lead to the same groupings, but they do have in common the separation of the centre and radial phenotypes. Manual classification is necessarily more diverse, since our aim was to group together cases that were broadly similar without considering the individual scores of each of the 10 lesions. It allows five distinct phenotypes to be separated, even though 20% of cases share common characteristics suggesting common pathophysiological mechanisms. Interestingly, FECDs with only small guttae that are never confluent (guttae only) constitute a rare (9.4%) and virtually pure group. It is highly unlikely that this phenotype is an early stage of a “centre” form, since patients were the same age and guttae cover the entire DM, whereas centre forms most often have a ring with few Guttae.

Our subgroup of 109 genotyped patients includes 84.4% of patients with TNR expansion (comparable to that reported by Wieben et al. [7]). To our knowledge, this is the first time that the absence of TNR expansion has been associated with a limited number of typical phenotypes: radial and/or fused guttae, absence of central PFL (i.e. with perfect visibility of guttae) with hypertrophy of the peripheral striae. This innovative description suggests that the response of CE to stress already identified in FECD pathogenesis varies significantly depending on their genetic background: enough to alter the formation and distribution of guttae but not enough to suppress guttae formation. The exact mechanism of deposition in the form of guttae rather than a uniform layer remains unclear.

This study has several limitations: 1/We have no clinical data and the stage of the disease at the time of transplantation may vary from one patient to another. A second prospective study is underway to analyse the clinical characteristics of our proposed histological subgroups (NCT05742321). 2/We did not study the cellular aspect for two reasons: the surgical trauma is significant and does not allow for a complete view of intact ECs, and we chose to focus first on ECM complexity. 3/Despite its relative transparency, some guttae buried in the PFL may remain invisible. 4/The PCA is not very effective, as the 4 PCs explain only 59% of the variance contained in the ten variables. This suggests that our interpretation was not sufficiently discriminating. We note, however, that the PCA has the merit of showing that there are variable groupings capable of separating very different forms of FECD. Automated image analysis methods should improve clustering. 5/ We have not yet investigated the much rarer mutations [37], nor the role of epigenetic modifications [38].

In summary, this large FECD series enabled us to describe new tissue structures and demonstrate the predominance of radially aligned structures, suggesting the existence of centripetal cell migration in their genesis. We propose a new histological classification into five distinct phenotypes, some of which exist only in the presence of TNR expansion (guttae only) and others are significantly more frequent when TNR are absent (fused guttae), suggesting different pathophysiological mechanisms.

## Data Availability

All data produced are available online at https://gofile.me/5SfuG/6jofA5FOf (password FECD500)

## Acknowledgements

We acknowledge the assistance of the technical team from the BiiO (C Perrache, S Urbaniak and JM Papillon). We would like to thank Mr Frédéric MASCARELLI, Centre de Recherche des Cordeliers, UMR S1138, University Paris Descartes, Paris, for his invaluable assistance in carrying out this work. The PhD Thesis of Ms Hanielle VAITINADAPOULE was funded by the Berthe FOUASSIER scholarship from the Fondation de France.

## Author contributions statement

GTh, JMP, ODC, SP and PG conceptualized and planned the experiments. GTh secured funding. HV and GTh conducted histopathology and scoring, supervised the project wrote, reviewed, and edited the manuscript. DO and RT performed genotyping. EC, EO and GT performed statistical analysis. HV, GTr, ODC, SP, ZH were responsible for tissue preparation and imaging. CM was in charge of image analysis and management of the online materials. HV and FF provided critical feedback and participated in manuscript review and editing. The FFSG provided the human tissue samples. All authors reviewed the manuscript, discussed the results, and contributed to its final version

## Conflict of interest statement

None related to the present paper.

## SUPPLEMENTARY MATERIAL ONLINE: figure legends

**Figure S1. Surgical Descemetorhexis.** Schematic representation of the surface of the Descemet membrane dissected during the 8 mm Descemetorhexis (in blue) in relation to the entire surface of the endothelium, i.e. approximately 45% of the total surface area (A). Intraoperative view of the Descemetorhexis at the stage when 1/3 of the diseased Descemet membrane has already been dissected (B).

**Figure S2. Assessment of pre-analytical conditions on the morphology observed with transmitted light optical microscopy.** Flat-mounted Descemet’s membrane from a Fuchs endothelial corneal dystrophy patient with one half fixed in 4% paraformaldehyde immediately after retrieval (PFA) versus the other half immersed in water immediately after retrieval and flat mounted after 10 weeks (H_2_O). The fixed piece presented multiple area with altered endothelial cells whereas the piece immersed in water appeared as completely decellularized.

**Figure S3. Representative examples of the three types of histological structures rarely described to date in Fuchs endothelial corneal dystrophy and used in our proposed classification.** Peripheral striae (A), curly fibres (B) and bands of guttae (arrow heads). Their analysis required observing each image at different magnifications.

## Notes

### Competing Interest Statement

The authors have declared no competing interest.

### Funding Statement

no funding

### Author Declarations

The handling of tissues adhered to the tenets of the Declaration of Helsinki of 1975 and its 1983 revision in protecting donor confidentiality and was approved by the ethics committee of the St-Etienne University Hospital (IRB_IORG0007394, Ref_IRBN1142021/CHUSTE).

