## Supplemental data for "Pathological classification of Fuchs endothelial corneal dystrophy in several types and their relationships with CTG18.1 expansion repeats"

Vaitinadapoulé et al.

**Supporting information**

**Influence of pre-analytical conditions**

We verified that the DM did not undergo morphological degradation during storage by comparing samples analysed on the same day with samples stored for between 1 and 52 weeks (Figure S2). In water (43% of cases) or balanced salt solution (BSS) (24% of cases), CECs disappeared but ECM morphology observed under optical microscopy remained intact. In paraformaldehyde (PFA) (33% of cases), residual cells persisted, but their morphology was often altered compared to healthy CECs (due to both the pathological process and surgical trauma (stripping and passage through a narrow incision)), which is why we quickly abandoned this method, as our objective was to analyse the ECM only. All 10 elementary lesions and 5 groups of the manual classification were observed under all 3 transport conditions. None of the lesions or groups disappeared after any of the 3 conditions. The distribution of the different lesions in the three environments was detailed in Table S1. For three of the ten items (Amor, Grad, Orad), the MDs in PFA had lower scores, suggesting that the fixed residual cells interfered with the observation of the finest structures (masking effect). Regarding peripheral striae, samples in BSS showed more striae, but these were larger DM feret’s diameter, Table S1) due to a centre effect (some ophthalmology centres producing larger Descemet striae were accidentally only supplied with collection tubes pre-filled with BSS).

By analysing the samples transported under these three conditions, we avoided bias (e.g. potential tissue shrinkage in a fixative or potential artefactual degradation in the absence of fixation), but we probably made reading more difficult (addition of noise), particularly due to residual cells that were fixed and distributed unevenly, which could cause certain details to be missed

**Table S1.** Influence of the pre-analytic medium on the reading of the 10 Descemet’s membrane characteristics.

| Criterion (abbreviation) | Grades | BSS | Water | PFA | P (Khi^2^) |
| --- | --- | --- | --- | --- | --- |
| Guttae number  (Gnum) | 0: free surface > surface covered by Guttae | 23,3% | 12,7% | 7,8% | P=0.002 |
|  | 1: free surface = surface covered by Guttae | 46,7% | 45,1% | 47,3% |  |
|  | 2: free surface < surface covered by Guttae | 30,0% | 42,3% | 44,9% |  |
| Guttae shape  (Gsha) | 0: only or mostly round | 85,0% | 89,7% | 83,8% | Not applicable |
|  | 1: balanced mixture of round and deformed Guttae | 15,0% | 9,9% | 13,2% |  |
|  | 2: only or mostly deformed | 0,0% | 0,5% | 3,0% |  |
| Guttae diameter  (Gdia) | 0: only or mostly small | 10,8% | 8,0% | 4,8% | Not applicable |
|  | 1: balanced mix of small and large diameters | 87,5% | 91,5% | 88,6% |  |
|  | 2: only or mostly large | 1,7% | 0,5% | 6,6% |  |
| Guttae overall pattern  (Gpat) | 0 centripetal gradient | 20,0% | 17,8% | 17,4% | Not applicable |
|  | 1 rich periphery | 6,7% | 3,8% | 6,0% |  |
|  | 2 homogeneous | 3,3% | 3,8% | 12,6% |  |
|  | 3 intermediate ring | 66,7% | 73,2% | 62,3% |  |
|  | 4 inclassable | 3,3% | 1,4% | 1,8% |  |
| Guttae radial orientation  (Grad) | 0: absent | 3,3% | 2,8% | 8,4% | P<0.001 |
|  | 1: present in 1 or 2 quadrants (=90 à 180°) | 30,0% | 18,3% | 35,3% |  |
|  | 2: present in more than 2 quadrants (>180°) | 66,7% | 78,9% | 56,3% |  |
| Guttae band  (Gban) | 0: absent | 90,0% | 83,6% | 91,0% | P=0.06 |
|  | 1: present | 10,0% | 16,4% | 9,0% |  |
| « Curly fibers »  (Curl) | 0: absent | 22,5% | 15,0% | 24,6% | P=0.153 |
|  | 1: Present without being the dominant structure | 65,8% | 74,6% | 67,1% |  |
|  | 2: Present on a massive scale | 11,7% | 10,3% | 8,4% |  |
| Amorphous extra cellular matrix  (Amor) | 0: absent | 16,7% | 16,0% | 6,6% | P=0.038 |
|  | 1: Present without being the dominant structure | 76,7% | 79,8% | 88,0% |  |
|  | 2: Present on a massive scale | 6,7% | 4,2% | 5,4% |  |
| Radial orientation of other structures (excluding Guttae)  (Orad) | 0: absent | 2,5% | 3,8% | 7,8% | P<0.001 |
|  | 1: present in 1 or 2 quadrants (=90 à 180°) | 15,8% | 16,0% | 35,3% |  |
|  | 2: present in more than 2 quadrants (>180°) | 81,7% | 80,3% | 56,9% |  |
| Peripheral striae  (Pstr) | 0: absent | 45,8% | 72,3% | 76,6% | P<0.001 |
|  | 1: present in 1 or 2 quadrants (=90 à 180°) | 35,0% | 19,2% | 17,4% |  |
|  | 2: present in more than 2 quadrants (>180°) | 19,2% | 8,5% | 6,0% |  |
| Feret min (mean+/-SD) | | 9.2+/-0.6 | 8.0+/-1.1 | 7.9+/-0.9 | P<0.001 |
| Feret max (mean+/-SD) | | 10.5+/-0.6 | 9.4+/-1.1 | 9.1+/-0.9 | P<0.001 |

**Methodological details for genotyping**

Blood samples were collected at the hospitals of Saint-Etienne and Metz. We used two techniques: Short Tandem Repeat Polymerase Chain Reaction (STR-PCR) and Tripled Primed Polymerase Chain Reaction (TP PCR) to quantify CTG18.1 TNR expansion in *TCF4* gene as previously described [1]. Genetic analyses were conducted at the Molecular Genetics Laboratory of the Saint-Étienne University Hospital (CHU). Based on the data from Wieben et al., we classified the genetic profiles into three groups: expansion-positive (>50 repeats), intermediate (40-50 repeats), and expansion-negative (<40 repeats) [2]. STR PCR is capable of precisely detecting up to 70-80 CTG18.1 TNR expansion, while TP PCR enables the detection of large-sized repeated alleles; however, beyond 100 repeats, it can only confirm the presence of a large allele without determining the exact number of repeats. The primers, PCR mixes, and cycle conditions followed Mootha et al. [1], as detailed in Tables S2, S3, and S4.

STR PCR involves amplifying a specific DNA sequence that contains a short nucleotide sequence repeated several times, using two primers flanking the repeat: P1 labelled with a fluorochrome (5' FAM) and P2 (**Table S2**). Capillary electrophoresis is then used to separate the fluorescent PCR products containing the repeat, allowing the identification of the repeat size in each allele. TP PCR also uses two primers flanking the repeat: P1 labeled with a fluorochrome (5' FAM) and P3, along with an additional internal primer P4 specific to the CTG18.1 repeat. The P4 primer has a 5' tail sequence corresponding to primer P3. This 5' tail of P4 has no homology with the human sequence (**Table S2**). P4 binds to multiple sites in the CTG repeat during the first amplification cycles, limiting the formation of secondary structures. Primer P3, which hybridizes to fragments containing primer P4, selectively continues the amplification of the obtained fragments. Capillary electrophoresis is then used to separate the fluorescent PCR products containing the repeat, allowing the identification of the repeat size in each allele. Genomic DNA was extracted from peripheral blood leukocytes, followed by a quality check. It was then diluted to a final solution with a concentration of 50 ng/µL. The primers used for STR PCR and TP PCR were those published by Mootha et al. and presented in **Table S2**[1]. PCR mixes were prepared with the Qiagen HotStarTaq DNA Polymerase kit, with the concentrations used for each mix listed in **Table S3**. The PCR cycles were performed using the parameters described by Mootha's team, shown in **Table S4**.

**Table S2: Primers for STR PCR and TP PCR**

| **Primer** | **Sequence 5’ – 3’** | **Genomic Position (GRCh38)** | **Technique** |
| --- | --- | --- | --- |
| P1  (5' FAM) | AATCCAAACCGCCTTCCAAGT | chr18:55,586,242-55,586,262 | STR PCR  TP PCR |
| P2 | CAAAACTTCCGAAAGCCATTTCT | chr18:55,586,076-55,586,098 | STR PCR |
| P3 | TACGCATCCCAGTTTGAGACG | No homology | TP PCR |
| P4 | TACGCATCCCAGTTTGAGACGCAGCAGCAGCAGCAG | 5’ tail – no homology  chr18:55,586,156-55,586,227 | TP PCR |

**Table S3: Composition of the mix used for STR PCR and TP PCR**

| **Reagent** | **STR PCR – Volume per Reaction** | **TP PCR – Volume per Reaction** |
| --- | --- | --- |
| Water | 10.5 µL | 7.6 µL |
| Buffer Q | 2.5 µL | 2.5 µL |
| dNTP (0.2 mM) | 2.5 µL | 2.5 µL |
| Solution Q | 5 µL | 5 µL |
| MgCl2 1.5 mM | 1.5 µL | - |
| Taq Q polymerase | 0.2 µL | 0.2 µL |
| Primer P1 (5' FAM) – F | 0.4 µL of a 0.3 µM solution | 1.2 µL of a 1 µM solution |
| Primer P2 – R (0.3 µM) | 0.4 µL | - |
| Primer P3 – R (1 µM) | - | 1.2 µL |
| Primer P4 – R (0.03 µM) | - | 0.8 µL |
| **DNA (50 ng/µL)** | **2 µL** | **4 µL** |
| **Total Volume** | **25 µL** | **25 µL** |

**Table S4: Parameters used for PCR Cycles**

|  | **Denaturation** | **Annealing** | | | | | | | **Extension** |
| --- | --- | --- | --- | --- | --- | --- | --- | --- | --- |
| **STR PCR** | 95°C 10 min | 94°C 30 sec | | | 58°C 30 sec | | | 72°C 30 sec | 72°C 10 min |
|  |  | 30 cycles | | | | | | |  |
| **TP PCR** | 95°C 9 min | 95°C 30 sec | 62°C 30 sec | 72°C 4 min | | 95°C 45 sec | 62°C 45 sec | 72°C 4 min  +15 sec of extension at each cycle | 72°C 10 min |
|  |  | 10 cycles | | | | 30 cycles | | |  |

The PCR products from both techniques were analysed by capillary electrophoresis on a SeqStudio (Applied Biosystems by Thermo Fisher Scientific) to separate the fluorescent fragments containing the repeat. For this, 0.5 µL of PCR products from one of the two techniques is added to 0.3 µL of GeneScan 500 Liz size marker and 14.2 µL of Hi-Di formamide. An additional denaturation step of 3 minutes at 86°C was performed for the TP PCR just before capillary electrophoresis migration. The results of the capillary electrophoresis were visualized using GeneMapper software to determine the size of the repeats (Figure S5).

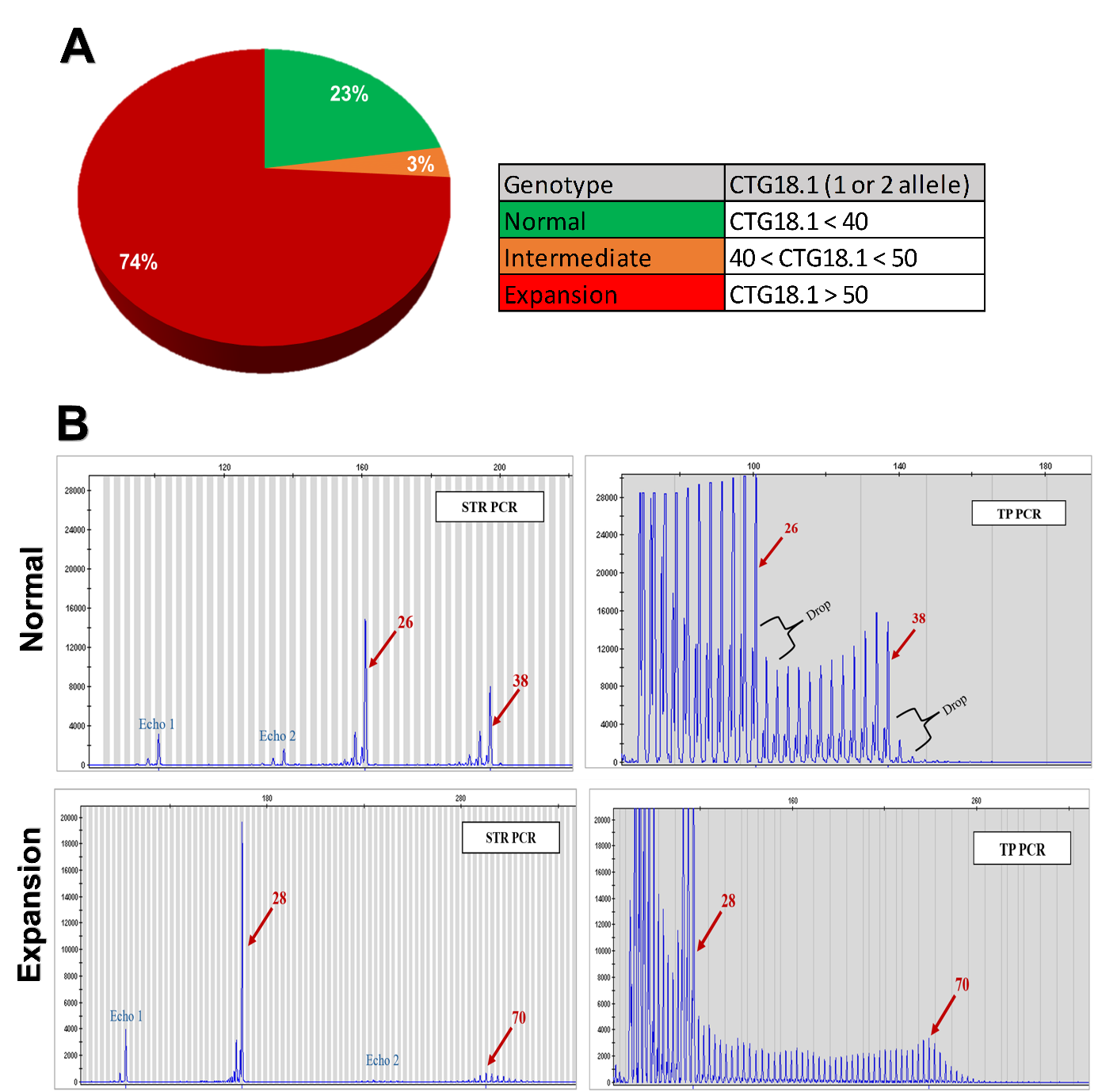

**Figure S4. Genotyping of CTG18.1 TNR expansion: Distribution of CTG18.1 expansion in the genotyped patients**: less than 40 repeats - normal allele; between 40 - 50 repeats - intermediate allele; more than 50 repeats - allele with CTG18.1 expansion (mutated) (A). **Results of CTG18.1 expansion analysis by STR-PCR and TP-PCR for representative genetic profiles** – 1) Normal heterozygote profile (at the top) - both alleles (26 and 38 repetitions) are visible using the two techniques. A systematic "echo" located 20 repeats upstream is observed for each allele in STR-PCR. In TP-PCR, the normal allele corresponds to the last peak before the "drop-off." 2) Mutated heterozygote profile (at the bottom) – the normal allele (28 repeats) is visible using the two techniques, but the second allele shows a broadened peak in STR-PCR. In TP-PCR, an expanded profile is detected by a characteristic train of peaks who confirm the pathological expansion and allows repeat sizing for the second allele (70 repeats) (B).

**The French Fuchs Study Group:**

Twenty-five Ophthalmology departments of the following hospitals (by alphabetic order or surgeons)

1. University Hospital of Grenoble: Dr Diane BERNHEIM, Pr Christophe CHIQUET
2. Hôpital National des 15-20, IHU ForeSight, GRC 32, Transplantation et Thérapies Innovantes de la Cornée, TTIC, INSERM-DGOS CIC 1423, Paris : Pr Vincent BORDERIE
3. University Hospital, Nouvel Hôpital civil, Strasbourg: Pr Tristan BOURCIER
4. University Hospital Cochin, Paris: Pr Jean-Louis BOURGES
5. University Hospital, Gabriel Montpied Clermont-Ferrand: Pr Frédéric CHIAMBARETTA
6. University Hospital Morvan, Brest: Pr Béatrice COCHENER
7. University Hospital of Dijon: Pr Catherine CREUZOT, Pr Louis ARNOULD, Dr Florian BAUDIN
8. University Hospital of Montpellier: Pr Vincent DAIEN
9. University Hospital, Robert Debré, Reims: Pr Alexandre DENOYER
10. University Hospital Sart Tilman, Liège, Belgium: Pr Bernard DUCHESNE
11. Kleber Ophthalmology Centre, Lyon: Dr Nicolas DUQUESNE
12. University Hospital, Purpan, Toulouse: Pr Pierre FOURNIE
13. University Hospital of Besançon: Pr Anne-Sophie GAUTHIER
14. University Hospital of Saint-Etienne: Pr Philippe GAIN, Pr Gilles THURET
15. Clinique Monticelli-Vélodrome Marseille: Pr Louis HOFFART
16. Ophthalmology Centre, Lausanne, Switzerland: Dr François MAJO
17. University Hospital Charles Nicolle, Rouen: Pr Marc MURAINE
18. Ophthalmology Centre, Valence, Dr Romain MOUCHEL
19. Hospital Center Metz-Thionville, Dr Jean Marc PERONE
20. University Hospital Caen/Normandie: Pr Jean Claude QUINTYN
21. Hospital Center, Antibes Juan les pins: Dr Alexandra RABOT
22. Rothschild Foundation Hospital, Paris: Dr Alain SAAD, Dr Damien GATINEL
23. Institut ophtalmologique de l’ouest, clinique Jules Verne, Nantes: Dr Pierre-Yves SANTIAGO, Dr Jean-Michel BOSC
24. University Hospital Pellegrin, Bordeaux: Pr David TOUBOULD
25. University Hospital, Hôtel Dieu, Nantes: Dr Bertrand VABRES
