## Supplementary figures and images for "Pathological classification of Fuchs endothelial corneal dystrophy in several types and their relationships with CTG18.1 expansion repeats"

### Supp Figure S1

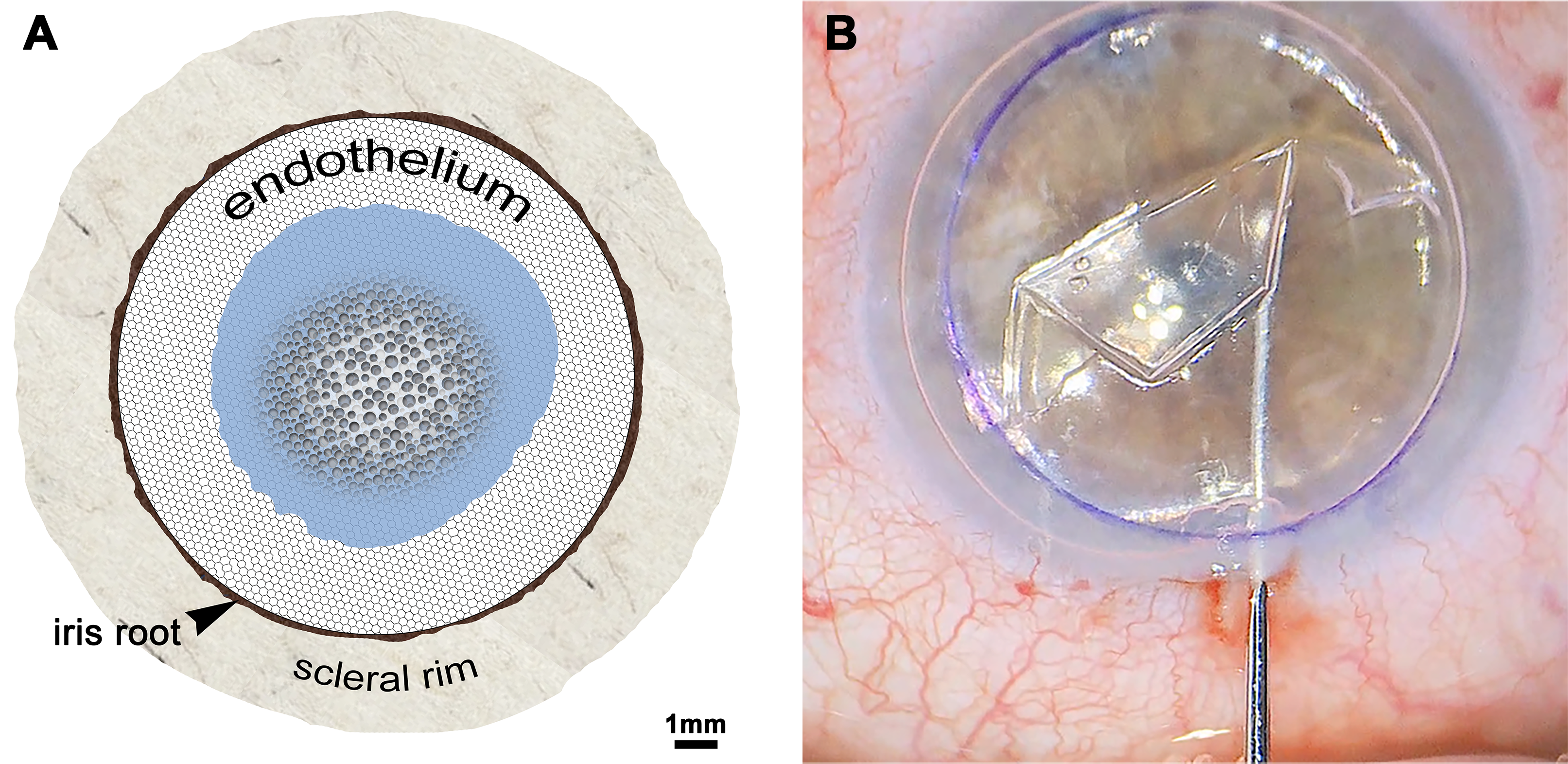

### Supp Figure S2

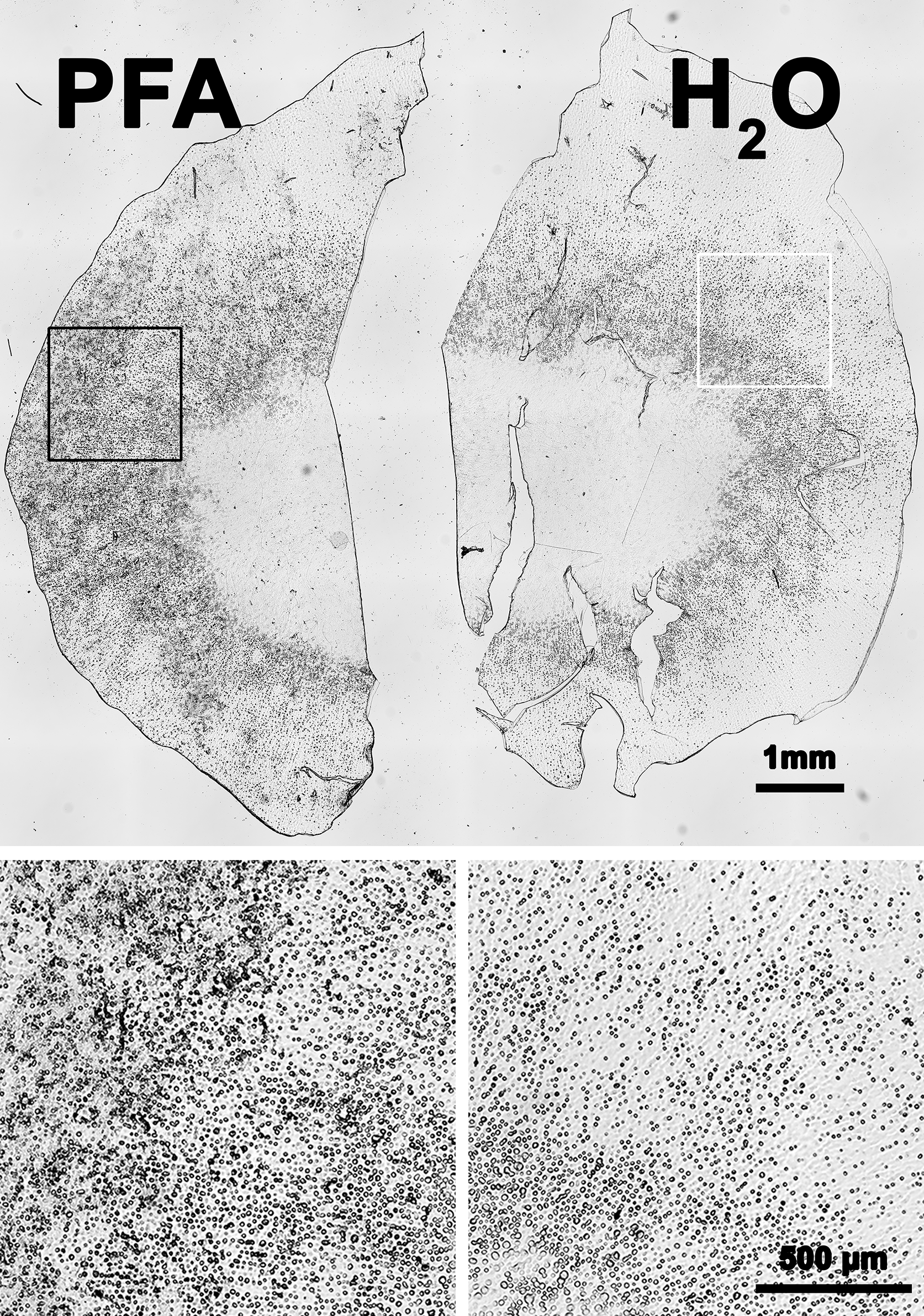

### Supp Figure S3

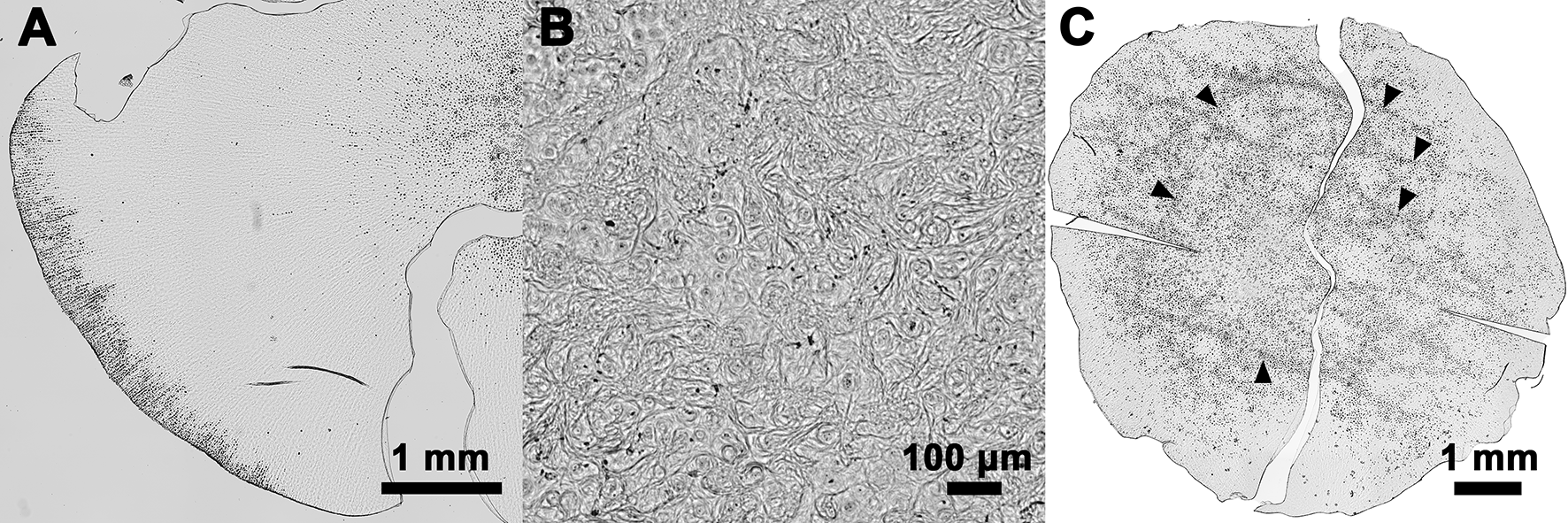
